# Art therapy is associated with a reduced rate of restrictive practices on an inpatient child and adolescent mental health unit

**DOI:** 10.1101/2022.12.13.22282997

**Authors:** Sarah Versitano, Artur Shvetcov, Joy Paton, Iain Perkes

## Abstract

**Background:** The elimination of seclusion and restraint, that is, restrictive practices, is a major aim of mental health services globally. The role of art therapy, a predominantly non-verbal mode of creative expression, is under-explored in this context.

**Aims:** To determine whether art therapy service provision was associated with a reduction in restrictive practices on an acute inpatient child and adolescent mental health services (CAMHS) unit.

**Methods:** The rate (events per 1,000 occupied bed days), frequency (percent of admitted care episodes with incident), duration, and total number of incidents of restrictive practices occurring between July 2015 – December 2021 were analysed in an ABAB design. The rate, frequency and number of incidents of intramuscular injected (IM) sedation, oral PRN (as-needed medication) use, and absconding incidents occurring in conjunction with an episode of seclusion or restraint were also analysed.

**Results:** The rate, frequency, duration, and total number of incidents of seclusion, the frequency and total number of incidents of physical restraint, and the rate, frequency and total number of incidents of IM sedation showed a statistically significant decrease during phases of art therapy service provision.

**Conclusions:** Art therapy service provision is associated with reduced use of restrictive practices in inpatient CAMHS.

## Introduction

The elimination of seclusion and restraint practices is a major aim of mental health services globally (Duke et al., 2014; Muir-Cochrane et al., 2014). Incidents of seclusion and restraint are associated with increased risk of psychological distress, trauma, humiliation, injury, and even death (Knox & Holloman, 2012; LeBel, et al., 2010; Nunno et al., 2006; Valenkamp et al., 2014). Seclusion is defined as an incident wherein a person is alone in an area which they are unable to freely exit, while restraint involves physical, chemical, or mechanical restriction of a person’s freedom of movement (Bureau of Health Information, 2019; NSW Health, 2020). The World Health Organisation (2019) has identified that these restrictive practices constitute a human rights violation, and are considered “cruel, inhuman, and degrading treatments” (p.28).

Furthermore, restrictive practices are associated with increased staff turnover, injury, absences, compensation claims, illness, and burnout (Bigwood & Crowe, 2008; Dean et al., 2010; LeBel & Goldstein 2005; LeBel et al., 2010). This indicates a significant impact on the wellbeing of mental health staff, who report an inadequate provision of viable alternatives to keep patients and staff safe (Bigwood & Crowe, 2008; Muir□Cochrane et al., 2018).

Inversely, a reduction in restrictive practices has been correlated with a shorter duration of hospital admission for patients, greater sustained success in the community after discharge (LeBel et al. 2010), fewer injuries to staff and young people, and a decrease in staff turnover (LeBel & Goldstein, 2005). When examining global approaches to reducing restrictive practices in inpatient care, there has been a move away from motivational programming, which includes behavioural reward systems, as this can in fact be counterproductive with young people in these settings, increasing the potential for seclusion or restraint to occur (Mohr et al., 2009). There has been an increased emphasis on prevention-oriented strategies and interventions to lower rates of restrictive practices in inpatient care. The literature on restrictive practices highlights the effectiveness of strategies which can improve self-regulation (Andrassy, 2016; Felver et al. 2017, Huckshorn, 2005; LeBel et al., 2004; NSW Health, 2020), interventions which are multimodal (Delaney, 2001; Gaskin et al., 2007; Huckshorn & LeBel, 2013), and those which integrate a person’s individualised interests (Caldwell et al., 2014).

The “Six core strategies to reduce seclusion and restraint use” developed by the National Association of State Mental Health Program Directors based in the USA (Huckshorn, 2004; Huckshorn et al., 2005) has been widely adopted in mental health settings (Azeem et al., 2018; Caldwell et al., 2014; Huckshorn 2004; 2005; Wieman et al., 2014). The fourth core strategy outlined in this framework relates to the use of seclusion and restraint prevention tools (Huckshorn, 2004). This strategy emphasises awareness of a person’s trauma history, and use of de-escalation tools including “creative changes to the physical environment, and daily, meaningful treatment activities” (p.28). No mention of art therapy as an effective prevention tool could be found in the literature on restrictive practices outlined above.

However, art therapy has the capacity to meet many of the aforementioned goals and recommendations, as it is a creative, prevention-oriented intervention which supports self-expression and self-regulation, and therefore has the capacity to mitigate escalating levels of distress.

### Art therapy in inpatient mental health services

The established benefits of art therapy in adult inpatient mental health services include developing greater self-awareness, social connection, self-expression, creativity, self-regulation, relaxation, and empowerment (Chiu et al., 2015; Dick, 2001; Laranjeira et al., 2019; Scope et al., 2017; Uttley et al. 2015). While the literature on child and adolescent mental health indicates similar benefits to adult cohorts, it tends to focus on either specific therapeutic interventions or techniques in an inpatient setting (Nielsen, 2018; Nielsen et al., 2019; Lyshak-Stelzer et al., 2007; Wyder, 2019) or broad approaches to art therapy in child and adolescent mental health (Case & Dalley, 2007; Malchiodi, 2015; Riley, 2001; Rubin, 2005). Although noting the limited empirical evidence for the use of art therapy with children in mental health services, a recent systematic review of the effectiveness of art therapy for children diagnosed with mental health disorders found benefits for children who had experienced trauma (Braito et al., 2021).

Concerningly, paediatric patients are far more likely to experience an incident of seclusion or restraint relative to adult patients (De Hert et al., 2011; NHS Benchmarking Network, 2019). Consumers of inpatient child and adolescent mental health services also have high rates of developmental trauma, and challenges with expressing their distress verbally, which can lead to an increased use of restrictive practices (Nielsen et al., 2019, p.165). Art therapy is a creative, psychotherapeutic intervention which allows young people to explore and process overwhelming experiences by identifying feelings non-verbally. Further to this, art therapy shows promise in reducing rates of restrictive practices for children and adolescents in inpatient mental health settings (Nielsen, 2018, p.4; Lyshak-Stelzer et al., 2007, p.167). In providing a safe, creative, non-verbal avenue of expression, levels of acute distress which often precipitate these incidents can be effectively de-escalated to achieve targeted prevention.

Our retrospective study examines the use of restrictive practices on an acute inpatient CAMHS unit in a metropolitan public hospital. The study aimed to identify whether there was an association between art therapy service provision, and a reduction in the use of restrictive practices. We examined over six years of data during which there were sustained periods of art therapy service provision which could be compared to periods without service.

## Materials and methods

Ethics approval for this study was granted from the Sydney Children’s Hospitals Network Human Research Ethics Committee (2021/ETH12228). A waiver of consent was also granted as this data had already been routinely collected by the service, remained de-identified for the purpose of the study, and was retrospectively accessed providing sufficient protection of privacy to participants.

Data on restrictive practices which occurred between July 2015 – December 2021 on an 8-bed acute inpatient CAMHS unit of a metropolitan public hospital in Sydney, Australia were extracted from two discrete sources in order to optimise validity and reliability. Data included incidents which were documented in the Incident Information Management System (IIMS), and incidents documented on paper in the seclusion and restraint registers stored on the inpatient unit. Data from these two sources were merged into a single data set, in order to generate the most accurate documentation of all incidents which occurred. Duplicates were discarded using the incident ID as a reference point. Data on intramuscular injected (IM) sedation, also referred to as chemical restraint when used to restrict a person’s movement (NSW Health, 2020), oral PRN use (i.e., as-needed medications), and absconding were also included if they occurred in immediate association with an incident of seclusion and/or physical restraint.

Data extracted from IIMS reports and the seclusion and restraint registers included the incident ID/IIMS number, date, time, patient identifier/medical record number (MRN), incident type, duration, history of trauma/abuse, reason for the use of restrictive practice/s, and a description of the incident. Incidents of seclusion and physical restraint were then analysed in a naturalistic ABAB design wherein phases of art therapy service provision were ‘B’, and phases without service provision were ‘A’ (Table 1.).

**Table 1.** Phases of service within naturalistic ABAB design.

| Phase | Date range | *n* months |
| --- | --- | --- |
| A - Pre-service | Jul 2015 - Dec 2017 | 30 |
| B - Initial service | Jan 2018 - Jan 2020 | 25 |
| A - Suspension of service | Feb 2020 – Oct 2020 | 9 |
| B - Reintroduction of service | Nov 2020 – Dec 2021 | 14 |

The art therapy service included both group and individual arts-based therapeutic interventions, integrating directive and non-directive approaches to art therapy, delivered by a masters-qualified registered art therapist. Young people were invited to engage in a weekly art psychotherapy group, a studio art therapy group, and could also be referred for individual art therapy sessions by the consultant psychiatrist and/or allied health case manager. The art psychotherapy group had a formal one-hour structure integrating a creative check in, artmaking time, a period of reflection on artworks and group process, and brief check out (Riley, 2001; Rubin, 2011). Studio art therapy groups were less formal in nature, running for 3 hours (generally during weekends), with flexibility around young people engaging with creative projects for brief periods, or the full duration of group (Moon & Lachman-Chapin, 2001). Individual sessions focussed on a range of therapeutic goals and were structured to meet the unique needs of the young person referred. Art therapy groups, and overall service coverage was originally scheduled on days and times when the Nurse Unit Manager had felt there were the highest levels of patient agitation, as well as inferences made about the common days and times when restrictive practices were occurring on the unit.

Demographic details were gathered from hospital medical records. MRN, age, sex, presenting issue, and primary diagnostic code were extracted for admissions occurring between July 2015 – December 2021. Data on occupied bed days, length of stay, number of admitted care episodes, and episode sequence were also extracted from the same source in order to calculate rate and frequency of incidents. Data on readmission rates were extracted from a New South Wales (NSW) Ministry of Health database for the period between January 2017 – December 2021 (readmission rates were not documented in this database prior to 2017). Data on length of stay and readmission rates were segmented into A and B phases, and a Mann-Whitney U Test was conducted to check for statistically significant differences between phases. Mean and standard deviation were also calculated for each phase.

Using the data set generated by the merged IIMS reports and seclusion and restraint registers, the rate, frequency, duration and total number of incidents of seclusion and physical restraint were calculated per month. The rate, frequency and total number of incidents of IM sedation, oral PRN use, and absconding were also calculated per month. Data was then segmented into the relevant A and B phases. All calculations were conducted using the same formulas as NSW Ministry of Health, replicating global standards for data reporting on restrictive practices (Bureau of Health Information, 2019; NSW Health, 2022). Existing NSW government formulas were used to calculate these variables in order to enhance the capacity to use implementation science for both clinical, and policy-based uptake of the research findings. Therefore, when calculating rate, this was the number of events per 1,000 occupied bed days. For frequency, this was the percent of acute mental health admitted care episodes wherein an incident occurred. For duration, this was calculated by averaging the length (in hours) of all incidents of seclusion and physical restraint occurring within a month.

The ABAB data set underwent a Shapiro-Wilk test to check for normality. Results indicated that non-parametric tests were appropriate for these data given the non-normal distribution of variables. Due to the naturalistic study design, phase subcomponents differed in length (range 9-27 months), however when both A phases (36 months) and B phases (38 months) were combined this resulted in similar time periods. To estimate similarity between restrictive practices occurring in the ABAB phases the Euclidean distance was calculated and a cluster dendrogram was built (Figure 1). Euclidean distance was calculated between all A and B datasets that included all variables of interest (i.e., the rate, frequency, duration and total number of incidents of restrictive practices). The cluster dendrogram showed that there are similarities in both A phases and B phases which provided justification for combining time periods. Therefore, a Mann-Whitney U test was undertaken to compare A phases (without service), with B phases (with service) to determine whether there were statistically significant differences. A Benjamini-Hochberg multiple correction was applied to adjust the *p* values and reduce the risk of a false positive. All inferential statistics were performed in Jasp (ver. 0.16.3.0).

**Figure 1.**
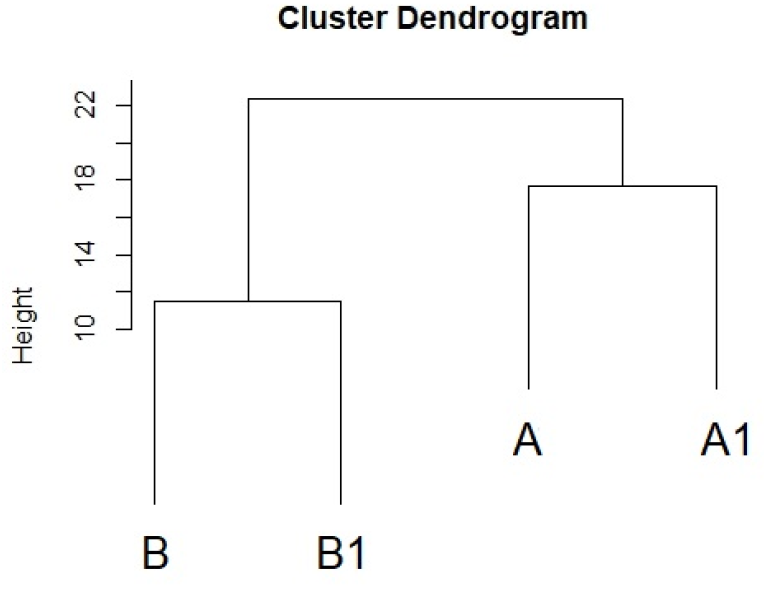
Similarity between A phases (without service) and B phases (with service).

Within the B phases (with service), incident dates were also examined to determine whether art therapy service delivery occurred on the same day as incidents of restrictive practices. Logistic regression was used to determine whether there were any statistically significant temporal associations with service delivery, as art therapy was offered on a part-time basis (3 days per week).

Diversional and therapeutic arts-based interventions were used sporadically on the unit prior to 2018, however no permanent art therapy service was established by an experienced art therapist prior to the initial B phase of service. There were also variations to nursing and medical staffing in addition to changes to the built environment throughout the study time-period. There was a 3-month gap in clinical data collection in 2016, and the unit was also closed for renovations in June 2019; both time periods were excluded from the analysis. There were also several waves of the covid-19 pandemic, however the service remained open throughout this time despite community lockdowns.

## Results

Between July 2015 and December 2021 there were 1,352 total admitted episodes of care in the acute inpatient CAMHS unit (258 males, and 1,094 females). Demographic information regarding young people who identified as gender fluid, transgender or non-binary was not extractable during this time-period, however clinical notes indicated that many young people admitted to the unit identified using names or pronouns which differed to those assigned at birth. The mean age of young people admitted to the unit was 14.1 years (SD = 1.65), and the average length of stay was 14 days (SD = 24.74).

Average length of stay during the A phases (without service) was 16.54 days (SD = 28.20), however during B phases (with service) there was a statistically significant decrease to an average of 11.59 days (SD = 20.72)(U = 261872, *p*-value = 0.00002). As outlined, data on readmission rates were only available for January 2017 – December 2021. Readmission rates for the A phases (without service) averaged 24.28% (SD = 10.62) and demonstrated a statistically significant decrease in B phases (with service) to an average of 16.5% (SD = 11.66) (U = 526, *p*-value = 0.03).

Young people were admitted to the unit with a range of primary diagnoses (Table 2.), predominantly mood (affective) disorders (25.59%), which comprised primarily of major depressive disorders (21.22%). Young people were also admitted due to suicidal ideation (13.68%), overdose (10.80%), anxiety disorders (10.50%), and eating disorders (8.65%).

**Table 2.**
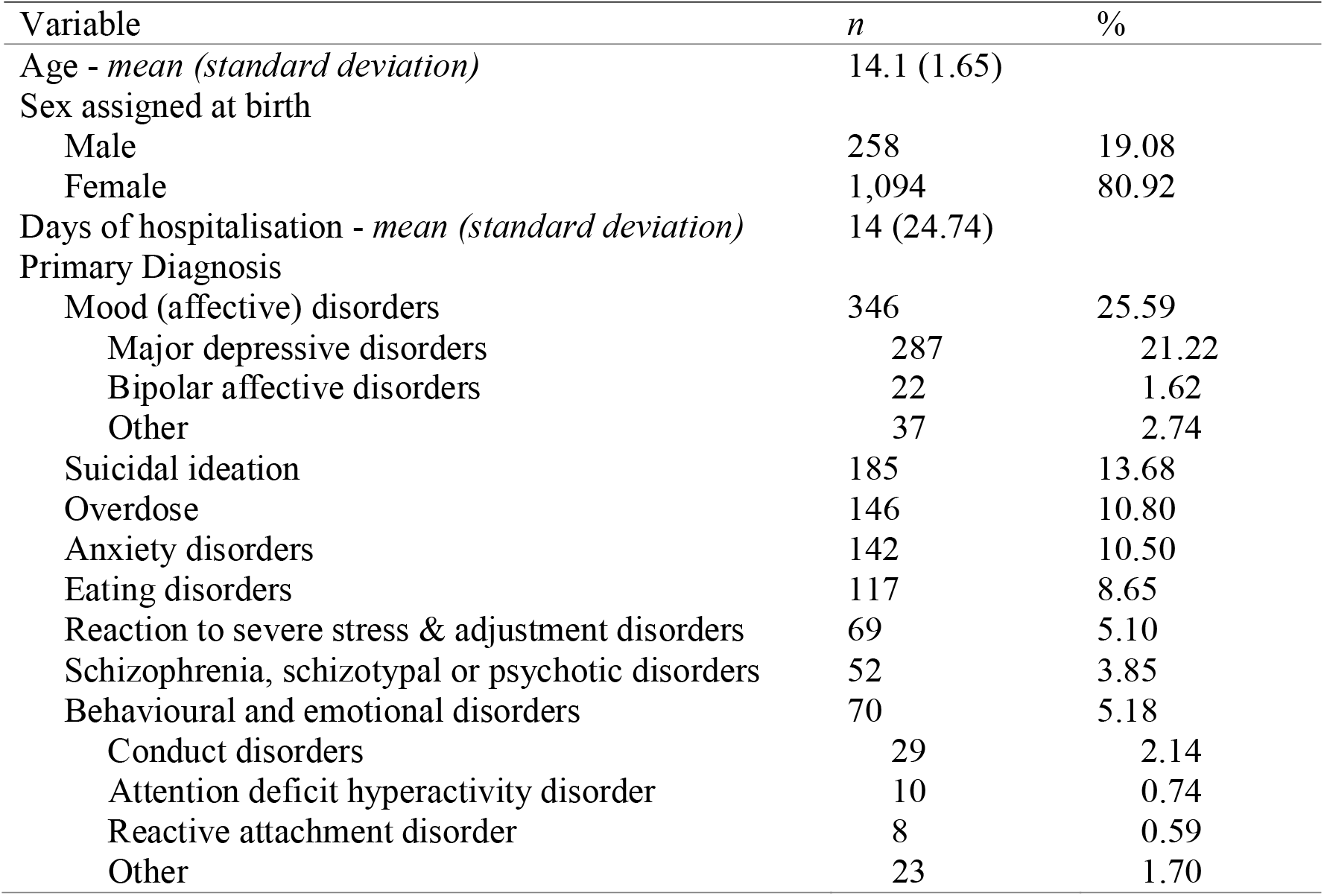
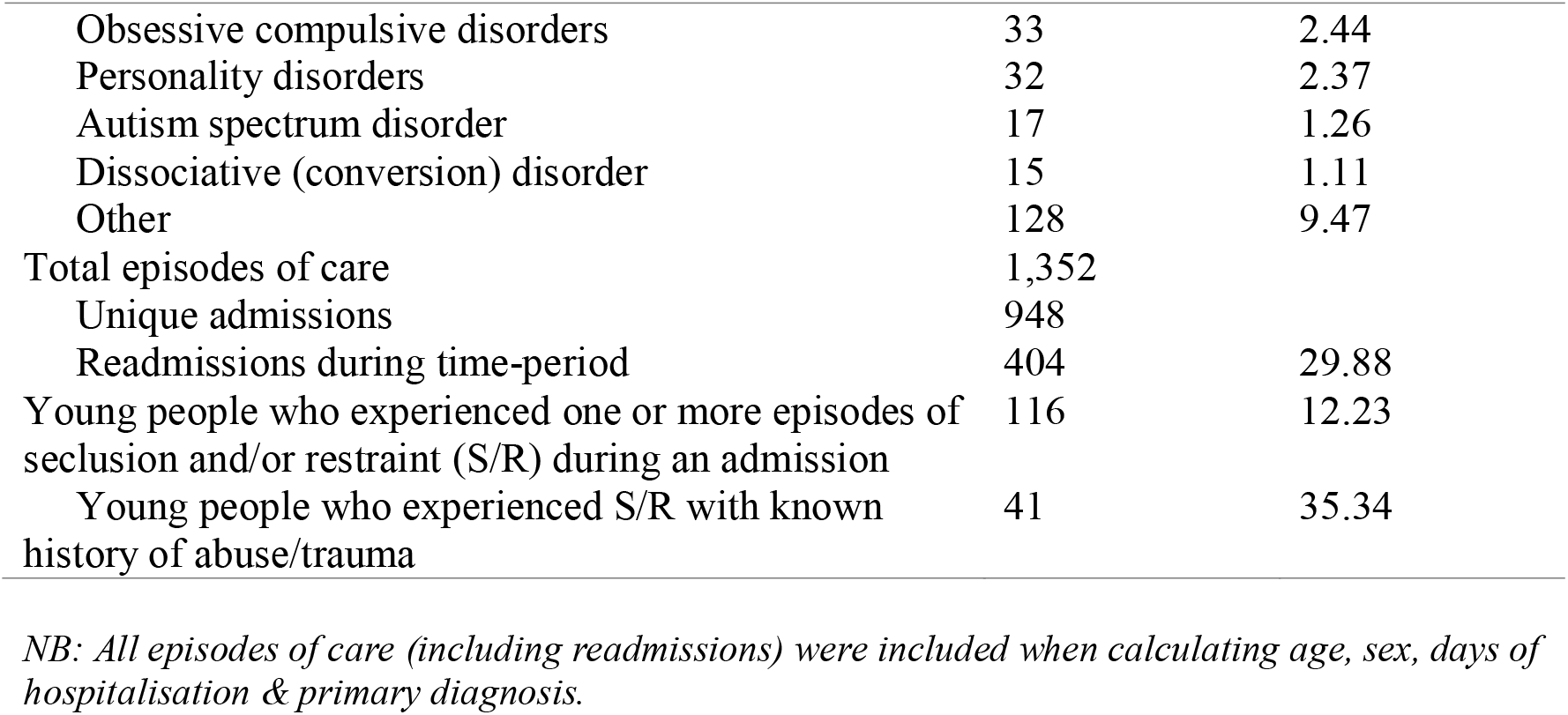
Clinical and demographic characteristics

Of the 1,352 episodes of care, 948 individual young people were admitted in this time-period. Of those 948 young people, 116 (12.23%) were involved in one or more incidents of seclusion and/or restraint during their time on the unit. Further to this, 41 (35.34%) young people who experienced an incident of seclusion and/or restraint also had a known history of abuse and/or trauma. There were 404 episodes of care (29.88%) which represent readmissions wherein a young person had one or more previous episodes of care on the unit (range 2-4 admissions).

For the episodes of care in this time-period, all variables (rate, frequency, duration and total number of incidents) relating to physical restraint, seclusion, IM sedation, oral PRN use and absconding incidents had a downward trend during B phases of art therapy service provision, when compared to A phases without service (Table 3. & Figure 2.).

**Table 3.** Associations between art therapy service provision and restrictive practices for A phases (without service) and B phases (with service).

| Variable | A phases<br>(without service) | B phases<br>(with service) | U Score | *p* value |
| --- | --- | --- | --- | --- |
| Months | 36 | 38 |  |  |
| Occupied bed days | 7892 | 7302 |  |  |
| Physical restraint |  |  |  |  |
| *n* incidents | 211 | 133 | 899 | 0.0370 |
| Rate | 26.74 | 18.21 | 871 | 0.0741 |
| Frequency | 22.57% | 11.57% | 909 | 0.0321 |
| Duration | 0.09 | 0.07 | 748 | 0.5960 |
| Seclusion |  |  |  |  |
| *n* incidents | 55 | 2 | 902 | 0.0027 |
| Rate | 6.97 | 0.27 | 899 | 0.0027 |
| Frequency | 5.68% | 0.34% | 903 | 0.0027 |
| Duration | 0.16 | 0.02 | 880 | 0.0048 |
| IM sedation |  |  |  |  |
| *n* incidents | 111 | 34 | 1047 | 0.0008 |
| Rate | 14.06 | 4.66 | 1022 | 0.0009 |
| Frequency | 13.62% | 3.30% | 1034 | 0.0008 |
| Oral PRN |  |  |  |  |
| *n* incidents | 69 | 39 | 737 | 0.2824 |
| Rate | 8.74 | 5.34 | 721 | 0.3546 |
| Frequency | 8.71% | 4.79% | 716 | 0.5072 |
| Absconding |  |  |  |  |
| *n* incidents | 34 | 26 | 800 | 0.5974 |
| Rate | 4.31 | 3.56 | 785 | 0.7010 |
| Frequency | 3.86% | 2.81% | 760 | 0.7014 |

*NB: Rate is calculated by events per 1,000 occupied bed days, frequency is calculated as the percentage of admitted care episodes with incident. Duration is averaged in hours. U score and *p*-value generated from Mann-Whitney U test.*

**Figure 2.**
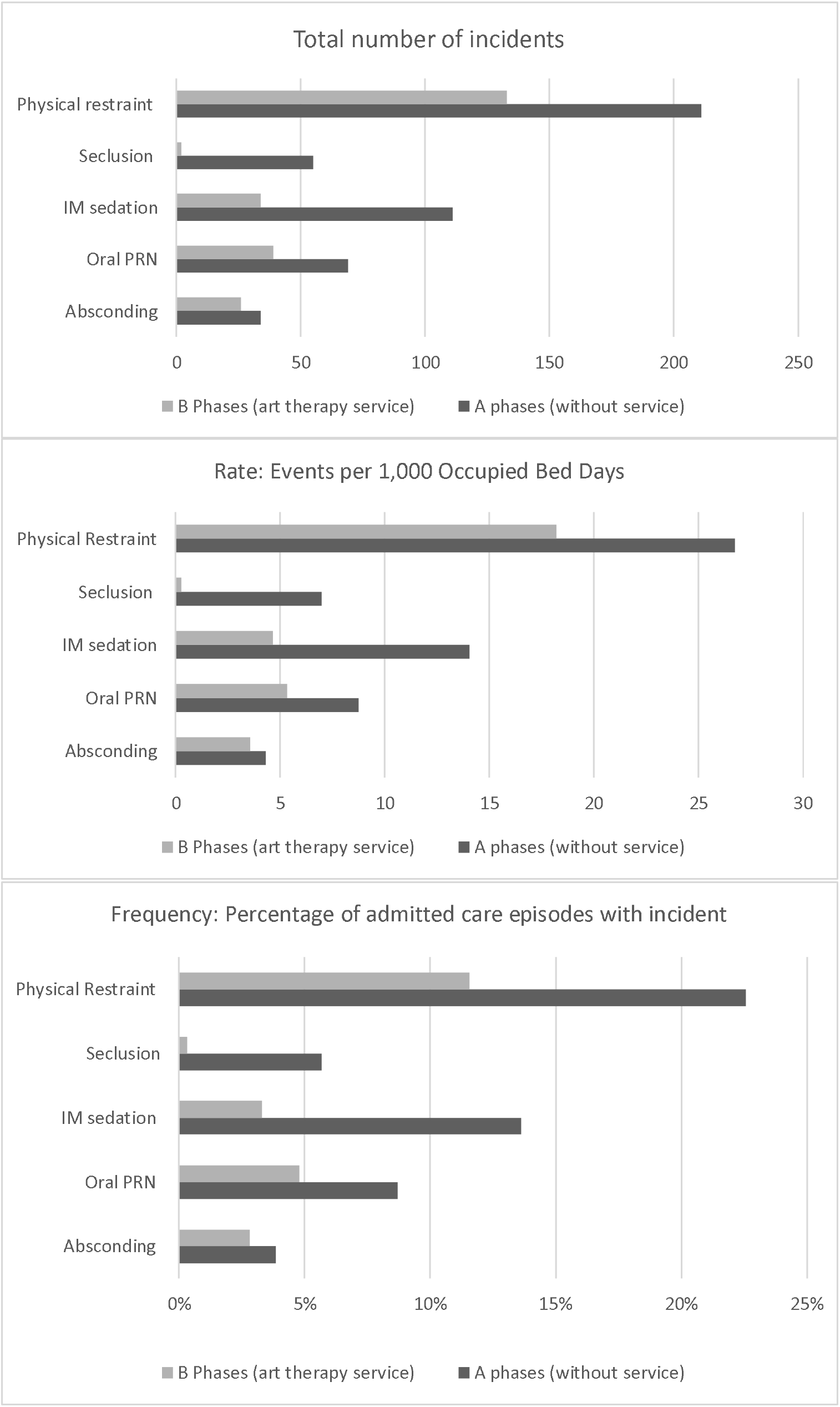
Total number of incidents, rate and frequency of restrictive practices occurring in A phases (without service) and B phases (with service).

Results of the Mann-Whitney U test comparing A phases (without service) to B phases (with service) showed a statistically significant decrease in the rate, frequency, duration and total number of incidents of seclusion during B phases of art therapy service provision. There was also a statistically significant decrease in the frequency and total number of incidents of physical restraint, and the rate, frequency and total number of incidents of IM sedation during B phases of art therapy service provision (U scores and *p*-values in Table 3.). Although there were clinically meaningful downward trends across all restrictive practices, the duration and rate of physical restraint, and variables relating to use of oral PRN and absconding incidents did not reach statistical significance between A and B phases.

When examining specific incident dates within B phases (with service), only 2 incidents of seclusion occurred, neither of which took place on the same day as the art therapy service. Within these B phases, the odds of physical restraint were also 49% lower on days with art therapy service provision when compared to days without (OR = 0.51, 95% CI [0.27, 0.91], *p*-value = 0.028).

## Discussion

The findings from this study indicate a clear association between the provision of art therapy, and a statistically significant reduction in the prevalence of restrictive practices such as seclusion, physical restraint, and IM sedation on an acute inpatient CAMHS unit. During phases without service provision on the unit, there are significantly higher rates of restrictive practices being used, indicating higher levels of acute distress for young people. When examining specific phases of art therapy service provision, no incidents of seclusion, and a statistically significant reduced likelihood of physical restraint were found during specific days when the art therapy service was present on the unit. Further to this, there was a statistically significant decrease in both the average length of stay, and average readmission rates during phases of art therapy service provision when compared to phases without service.

Higher readmission rates and length of stay indicate increased time periods wherein a young person remains in a restrictive, locked hospital environment, while also increasing costs associated with this level of care (Feng at al., 2017). While shorter length of stay has been associated with increased readmission rates (Figueroa et al., 2004), our analysis demonstrated a significant reduction in both these measures during phases of art therapy service provision. From this, it can be inferred that engagement with art therapy during an inpatient admission can decrease the need for an extended length of stay, while also providing positive effects which can extend beyond discharge, therefore decreasing the likelihood of readmission.

For some young people, the artworks created during an admission may provide visual evidence of their change or progress over time (Figure 3.). During the final closure session with the art therapist, a young person can reflect upon the body of work that has been created, and form this into a clear, integrated narrative of their personal journey during an admission. In doing so, their perceptions of change can be sustained after therapy concludes, as artworks can remain with the young person long after discharge. As Hanes (2001) identifies, the permanence of the artwork produced in sessions is unique to art therapy when compared to verbal therapies and provides “tangible and undeniable testimony to their progress” (p.159).

**Figure 3.**
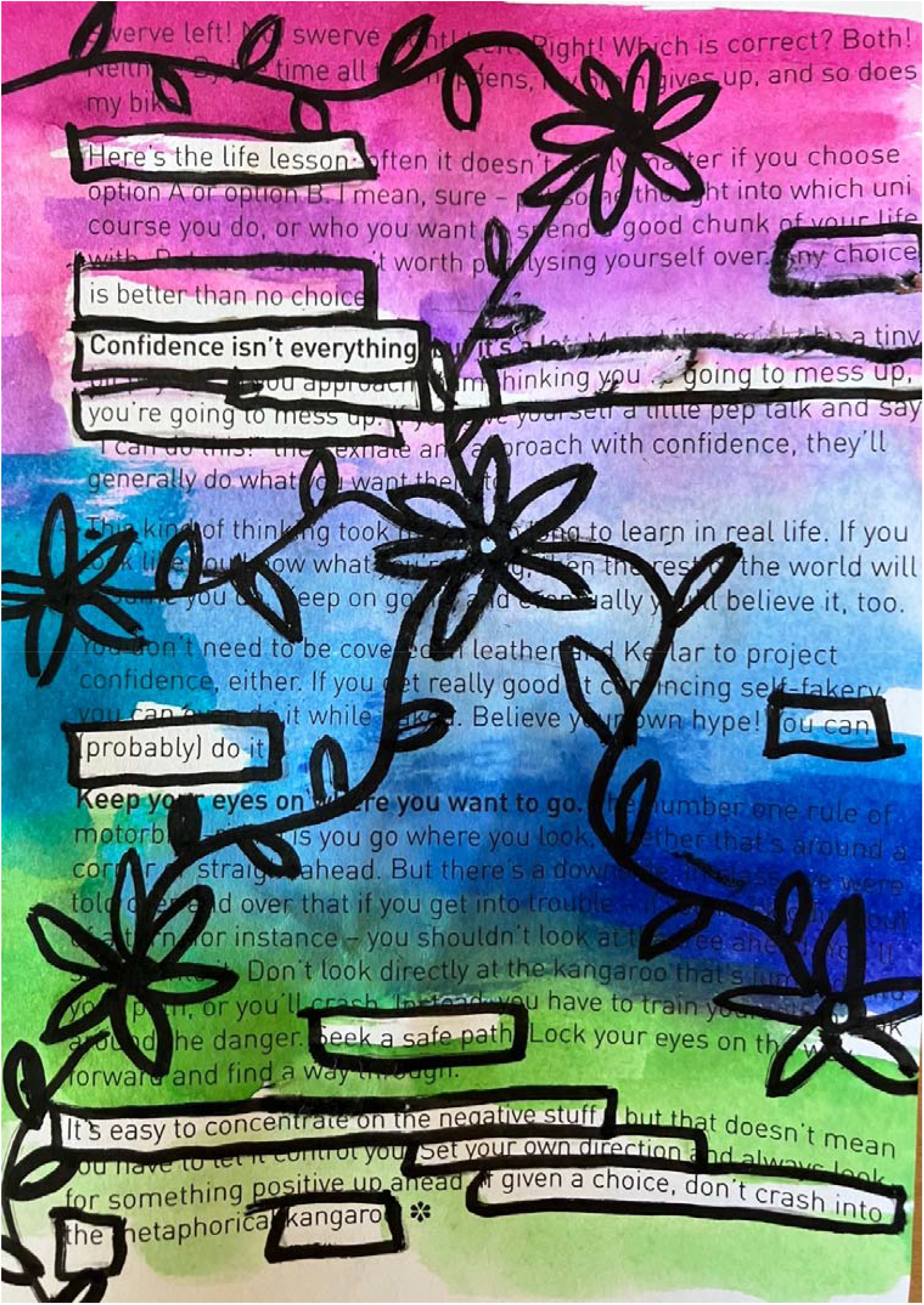
Artwork created by young person during final art psychotherapy group prior to discharge. NB: Erasure poem, a form of found poetry, created with mixed media “Here’s the life lesson… any choice is better than no choice…confidence isn’t everything… if you approach thinking you’re going to mess up, you’re going to mess up… You can (probably) do it… You can… seek a safe path… It’s easy to concentrate on the negative stuff… set your own direction… if given a choice, don’t crash into the kangaroo”

The art therapy service was thoughtfully integrated into the existing therapeutic program on the unit. Art therapy groups and overall service coverage was initially scheduled on particular days when the Nurse Unit Manager had noticed increased levels of agitation and boredom for young people, as well as inferences made from existing reports in the seclusion and restraint registers regarding the most common days and times when restrictive practices were occurring. Young people attended a weekly art psychotherapy group as well as a studio art therapy group. The art psychotherapy group encouraged self-expression, creativity, socialisation, interpersonal learning, and a therapeutic group culture wherein participants had a safe space to share their thoughts, feelings, and experiences (Riley, 2001; Rubin, 2011). The studio art therapy group also provided a safe, therapeutic space for young people to engage with independent creative projects with the presence and support of the art therapist (Moon & Lachman-Chapin, 2001). Both groups provided opportunities for social engagement, self-expression, and emotional regulation.

Boredom, which can be a result of occupational deprivation commonly occurring in restrictive environments such as a locked unit (Whiteford et al., 2020), often precipitates increased agitation or aggression, and is found to be a contributing factor in the potential increase of restrictive practices (Larue et al., 2009, NSW Health, 2017). The presence of a multidisciplinary team with the skills and capacity to provide trauma-informed, recovery-oriented care, and a predictable therapeutic program has been recommended to address this risk (NSW Health, 2017, p.41). Providing creative therapeutic services such as art therapy which can effectively engage young people, support emotional regulation, and encourage positive social interaction may therefore contribute to the prevention of precipitating factors such as aggression or agitation which typically increase prior to an incident of seclusion and/or restraint.

Our findings provide evidence for the importance of establishing creative therapeutic modalities and interventions in acute inpatient mental health care settings. Specifically, the provision of permanent art therapy services run by qualified registered art therapists in being able to support the reduction and prevention of harmful restrictive practices for children and adolescents. Young people who are admitted for care in an acute inpatient mental health setting have often trialled traditional psychotherapeutic and psychopharmacological interventions within a community setting and found these treatments to be ineffective in mitigating their mental health challenges (NSW Health, 2011; Sydney Children’s Hospitals Network, 2020). Creative, person-centred interventions such as art therapy provide a safe, inviting alternative for young people to engage with during an inpatient admission.

The promising findings of this study warrant further investigation into consumer perspectives (Springham & Xenophontes, 2021), particularly insights into their experiences of restrictive practices within these settings (Duke et al. 2014). Further research into consumer perspectives on art therapy within acute inpatient CAMHS services is currently being undertaken by the authors. This research aims to establish a deeper understanding of young people’s experience of art therapy as an intervention which can support emotional regulation, and therefore reduce the prevalence of restrictive practices in inpatient care.

When examining successful seclusion and restraint prevention programs for young people (Caldwell et al., 2014), a consumer from the Youth Development Institute in the United States offered the following advice to services aiming to eliminate restrictive practices:

> Be patient, and talk to us like people…Listening to us instead of telling us what to do could have stopped many restraints…Skilled staff don’t feel like they have to control us…they adapt and still keep us safe. (p.34)

This advice provides invaluable insights into the importance of a client-centred approach which focusses on deep listening, patience, and flexibility. Art therapy does just this, offering an alternative to utilising restrictive practices which can be experienced as controlling, punitive and traumatising (Caldwell et al., 2014; Knox & Holloman, 2012). More than one third of young people who had experienced an incident of seclusion and/or restraint in our study had a known history of abuse and/or trauma. Trauma-informed care emphasises the need for psychological, emotional, and physical safety to provide opportunities for a sense of control and empowerment (Hopper et al., 2010), both of which are removed when restrictive practices are enacted.

As outlined, young people on the acute inpatient CAMHS unit in our study could also be referred for individual art therapy by the treating team. These referrals were made for a variety of reasons. It may have been that visual or symbolic exploration was considered a gentler avenue of expression, e.g., when a young person has a known history of trauma, or perhaps a young person identified that art making was an existing therapeutic strategy, and the treating team wished to explore therapeutic goals through a creative lens. A young person may also struggle to engage effectively with verbal psychotherapies, perhaps due to challenging experiences with past clinicians, selective mutism, or severe psychotic symptoms. Art therapy is an adaptive therapeutic intervention which provides a safe space for a young person to express themselves creatively with flexibility around any need for verbal reflection. Trauma-informed art therapy practices acknowledge that verbal processing can often be challenging to access at times of increased distress, and that the artmaking can be an emotionally regulating experience when conducted within a safe, therapeutic relationship with a trained professional art therapist (Chong, 2015; Nielsen, 2018).

Nielsen (2018), in her work with children and adolescents in a high severity long-stay adolescent inpatient setting in Australia, identified that using art making in a therapeutic relationship was an effective de-escalation strategy, and preventative alternative to the use of restraint or seclusion when young people were struggling with aggressive behaviours on the unit. One young person on the unit shared that art therapy was “one of the only ways I could communicate the pain I was going through” (Nielsen et al., 2019, p.168). In a study examining the efficacy of trauma-focused art therapy in reducing post-traumatic stress disorder (PTSD) symptoms in an inpatient psychiatric facility for youth (Lyshak-Stelzer et al. 2007), findings showed that there was substantially more PTSD symptom reduction for the trauma-focused art therapy group when compared with the control group. Further to this, there was a reduced likelihood of incidents of seclusion occurring for young people in the trauma-focused art therapy condition (p.167). These findings, in addition to our study, provide evidence to support the value of art therapy interventions in supporting the reduction of restrictive practices with children and adolescents in an inpatient mental health setting.

It is also important to acknowledge the complexity of the therapeutic environment on an acute inpatient unit, and the varying factors, e.g., staffing changes, built environment variations, and the pandemic, which can influence an increase or decrease in restrictive practices. Nonetheless, despite a significant increase in the number of self-harm or suicidal ideation presentations in children and adolescents to emergency departments in NSW, Australia since 2015 (Sara et al., 2022), restrictive practices still significantly declined on this unit after the introduction of the art therapy service.

## Conclusion

While the eventual elimination of restrictive practices in inpatient mental health services is a multi-faceted and ongoing endeavour, art therapy can effectively and significantly contribute alongside other systemic and therapeutic interventions to the reduction of these harmful practices. Our study has established that there was a statistically significant reduction in incidents of seclusion, physical restraint, and intramuscular injected sedation during the phases wherein the art therapy service was present on the acute inpatient CAMHS unit.

Furthermore, there was a significant reduction in length of stay and readmission rates when art therapy was provided. These compelling findings are relevant for mental health services internationally, especially given the scarcity of literature on art therapy in child and adolescent mental health care (Braito, 2021; Cornish, 2013).

Although art therapy is not a panacea for the elimination of restrictive practices in inpatient mental health services, this study has demonstrated it can significantly contribute to the reduction of such practices. This has positive implications for the wellbeing of both young people and the staff providing care, and for reducing both the human and financial cost of restrictive practices. Given this, consistent and permanent art therapy programs run by qualified art therapists should be a core activity in all child and adolescent mental health services. Reducing restrictive practices in these settings is certainly a complex undertaking, however art therapy has an important role in providing effective, trauma-informed support to young people in acute inpatient care.

## Data Availability

All data produced in the present study are available upon reasonable request to the authors

## Acknowledgements

The authors acknowledge the Health Education and Training Institute (HETI) who provided funding to undertake this research as part of the Mental Health Research Award for 2022/23. The young people whose experiences were able to provide greater insights into how to effectively reduce these harmful practices. The nursing, allied health and medical staff on the inpatient CAMHS unit who work tirelessly to create a safe, therapeutic environment for young people. Kim Gregorio & Dr Sheridan Linnell who provided their expert guidance and support.

## Declaration of interest statement

The first author acknowledges her dual role as both clinician and researcher. No further conflicts of interest have arisen.

## Notes

### Competing Interest Statement

The authors have declared no competing interest.

### Funding Statement

This study was partially funded by the Health Education and Training Institute (HETI) Mental Health Research Award.

### Author Declarations

Ethics approval for this study was granted from the Sydney Childrens Hospitals Network Human Research Ethics Committee (2021/ETH12228). A waiver of consent was also granted as this data had already been routinely collected by the service remained de-identified for the purpose of the study and was retrospectively accessed providing sufficient protection of privacy to participants.

